# Prevalence and Characterization of Bacteria Present in the Nasopharynx of Outpatients with and without SARS-CoV-2

**DOI:** 10.1101/2022.08.17.22278915

**Authors:** James F. Shurko, Chris A. Mares, Robert B. Page, Kristina Lopez, Vivian Nguyen, Niti Vanee, Pramod K. Mishra

**Affiliations:** iGenomeDx, San Antonio, Texas, USA; Department of Life Sciences, Texas A&M University-San Antonio, San Antonio, Texas, USA

## Abstract

COVID-19 has emerged as a highly contagious and debilitating disease caused by the SARS-CoV-2 virus and has claimed the lives of over 6.8 million people worldwide. Bacterial co-infections are one of many co-morbidities that have been suggested to impact the outcome of COVID-19 in patients. The primary goal of this study was to assess the prevalence of bacterial co-infections and to describe any trends observed during the height of the COVID-19 pandemic. To do this, we investigated SARS-CoV-2 and bacterial co-infections from outpatient RT-PCR testing in Texas. The results indicate *Staphylococcus aureus, Streptococcus pneumoniae, Klebsiella pneumoniae, Moraxella catarrhalis*, and *Haemophilus influenzae* were the most frequently detected bacterial pathogens in both SARS-CoV-2 positive and SARS-CoV-2 negative patients and that these bacterial pathogens were present in these two patient populations at similar proportions. We also detected *Staphylococcus aureus* in a significantly larger proportion of males relative to females and people under 65 years of age relative to those 65 and over. Finally, we found that Hispanics were 75% more likely to be SARS-CoV-2 positive than non-Hispanics. The results suggest that COVID-19 patients may benefit from rapid diagnostic tests for bacterial pathogens and that this information could help delineate targeted antimicrobial therapy.

## Introduction

Coronavirus disease 2019 (COVID-19) is caused by Severe Acute Respiratory Syndrome Coronavirus 2 (SARS-CoV-2) and represents an important global health challenge. Approximately 6.8 million deaths have been reported as due to COVID-19 globally with over one million occurring in the United States^1,2^. In the state of Texas alone, the CDC has reported a total of over 6.8 million cases and over 86 thousand deaths since March of 2020. As of May 26^th^, 2022, the CDC reports an incidence of over 4.5 thousand cases per day contributing to an average of 5 daily deaths^3^. While vaccines are available to the general public in the United States, nearly 33.4% of the general population is not fully vaccinated (defined as receiving two doses of either the Pfizer or Moderna vaccine or a single Dose of the Johnson and Johnson vaccine) and in the State of Texas, approximately 34.4% of the population is not fully vaccinated^4,5^. These data, combined with the discovery of newly emerging strains, demonstrate that COVID-19 is a continuing health concern^6^.

SARS-CoV-2 infections display a wide range of prognoses ranging from asymptomatic infection to severe respiratory failure and death. More specifically, patients may experience fever, cough, fatigue, dyspnea, diarrhea, loss of taste or smell, aches, conjunctivitis, pneumonia, acute respiratory distress syndrome (ARDS) and multi-organ failure^7,8^. Currently, treatments for COVID-19 are limited and controversial, with clinicians focusing largely on supportive care^9-11^. In addition to comorbidities such as hypertension and diabetes, bacterial co-infections have been shown to have a profound impact on the outcomes of patients infected with SARS-CoV-2^12-15^. Subsequently, empiric antimicrobial therapy has been commonly administered to affected patients. Widespread antibiotic use remains controversial however, due to increased healthcare costs, increased adverse drug reactions, and the evolution of antimicrobial resistance. This study therefore aims to determine the effect of patient demographics (age, sex, race and ethnicity) on the frequency of SARS-CoV-2 and bacterial infection, and to compare the prevalence of common respiratory pathogens present within the nasopharynx of SARS-CoV-2 negative and SARS-CoV-2 positive patients. By elucidating the types of bacteria, frequency of bacterial co-infection, and populations most likely to be infected, these data may aid in guiding the selection of appropriate antimicrobials during infection as well as identify populations that are most likely to benefit from concurrent respiratory pathogen testing or antibiotic use.

## Results

Of the 4905 patients included in this data set 985 tested positive for SARS-CoV-2. Table 1 shows point estimates and 95% confidence intervals for the proportion of patients from various demographics (i.e., sex, age, ethnicity, and race) that were positive for SARS-CoV-2, S. *aureus*, and bacterial pathogens other than *S. aureus*. Of the 12 tests conducted to compare positivity across these demographics, three were statistically significant after correcting for multiple testing (Supplemental File 1). Despite fewer than half (46.20%) of all patients self-reporting ethnicity, the positivity rate for SARS-CoV-2 was 75% higher in Hispanics compared to non-Hispanics (Table 1). Further, the positivity rate for *S. aureus* was 23% higher in males than females and 24% higher in patients under 65 years of age when compared to individuals 65 years of age or older (Table 1). Finally, there were no statistically significant differences in the positivity rates of non-*S. aureus* bacteria detected across any of the demographic variables examined.

**Table 1.** Proportion of patients who tested positive for SARS-CoV-2 or respiratory pathogens stratified by demographic.

| Demographic | Number of Patients | Proportion (95% Confidence interval) |  |  |
| --- | --- | --- | --- | --- |
|  | n | SARS-CoV-2 | *S. aureus* | Other<sup>a</sup> |
| Sex |  |  |  |  |
| Female | 2837 | 0.1939 (0.1795-0.2089) | **0.2330 (0.2175-0.2490)** | 0.0917 (0.08128-0.1029) |
| Male | 2014 | 0.2105 (0.1929-0.2290) | **0.2855 (0.2659-0.3058)** | 0.1033 (0.0903-0.1174) |
| Undisclosed sex | 54 | 0.2037 (0.1063-0.3353) | 0.2037 (0.1063-0.3353) | 0.0556 (0.0116-0.1539) |
| Age |  |  |  |  |
| ≥ 65 years of age | 846 | 0.2163 (0.1890-0.2456) | **0.2116 (0.1845-0.2407)** | 0.0780 (0.0609-0.0982) |
| < 65 years of age | 4037 | 0.1982 (0.1860-0.2108) | **0.2631 (0.2495-0.2769)** | 0.0991 (0.0900-0.1087) |
| Undisclosed age | 22 | 0.0909 (0.0112-0.2916) | 0.2727 (0.1073-0.5022) | 0.2273 (0.0782-0.4537) |
| Race |  |  |  |  |
| African American | 65 | 0.1846 (0.0992-0.3003) | 0.2769 (0.1731-0.4019) | 0.1077 (0.0444-0.2094) |
| Asian | 36 | 0.1389 (0.0467-0.2950) | 0.3056 (0.1635-0.4811) | 0.0556 (0.0068-0.1866) |
| Native American | 7 | 0.1429 (0.0036-0.5787) | 0.2857 (0.0367-0.7096) | 0.2857 (0.0367-0.7096) |
| Pacific Islander | 1 | 0.0000 (0.0000-0.9750) | 0.0000 (0.0000-0.9750) | 0.0000 (0.0000-0.9750) |
| White | 1892 | 0.2378 (0.2188-0.2577) | 0.2775 (0.2574-0.2983) | 0.0946 (0.0818-0.1087) |
| Undisclosed race | 2904 | 0.1780 (0.1643-0.1924) | 0.2379 (0.2226-0.2539) | 0.0968 (0.0863-0.1081) |
| Ethnicity |  |  |  |  |
| Hispanic | 1983 | **0.2532 (0.2341-0.2729)** | 0.2552 (0.2361-0.2750) | 0.0872 (0.7519-0.1005) |
| Non-Hispanic | 283 | **0.1449 (0.1060-0.1914)** | 0.3145 (0.2608-0.3721) | 0.1060 (0.0727-0.1479) |
| Undisclosed ethnicity | 2639 | **0.1675 (0.1534-0.1823)** | 0.2471 (0.2307-0.2640) | 0.1016 (0.0903-0.1137) |
| Total | 4905 |  |  |  |

Proportions and 95% confidence intervals (95% CI) of patients with SARS-CoV-2, *S. aureus* and
(a) other bacteria contained in the respiratory testing panel regardless of *S. aureus* presence.
Statistically significant findings are bolded.

A total of eight bacterial pathogens were detected in this study including *S. aureus* (*n* = 1247), *S. pneumoniae* (n = 184), *M. catarrhalis* (n = 157), *K. pneumoniae* (n = 129), *H. influenzae* (n = 91), *M. pneumoniae* (n = 3), *C. pneumoniae* (*n* = 4) and *Bordetella sp (n* = 4). The estimated proportions and 95% confidence intervals of SARS-CoV-2 positive and SARS-CoV-2 negative patients who tested positive for each of these eight bacterial pathogens are shown in Figure 1. Three of these eight bacterial pathogens (*Bordetella sp*., *C. pneumoniae, and M. pneumoniae)* were detected in fewer than five patients; therefore, tests of equal proportions were not performed. Of the remaining five pathogens, tests comparing positivity rates in SARS-CoV-2 negative and SARS-CoV-2 positive patients were not statistically significant after correcting for multiple testing (Supplemental File 1).

**Figure 1:**
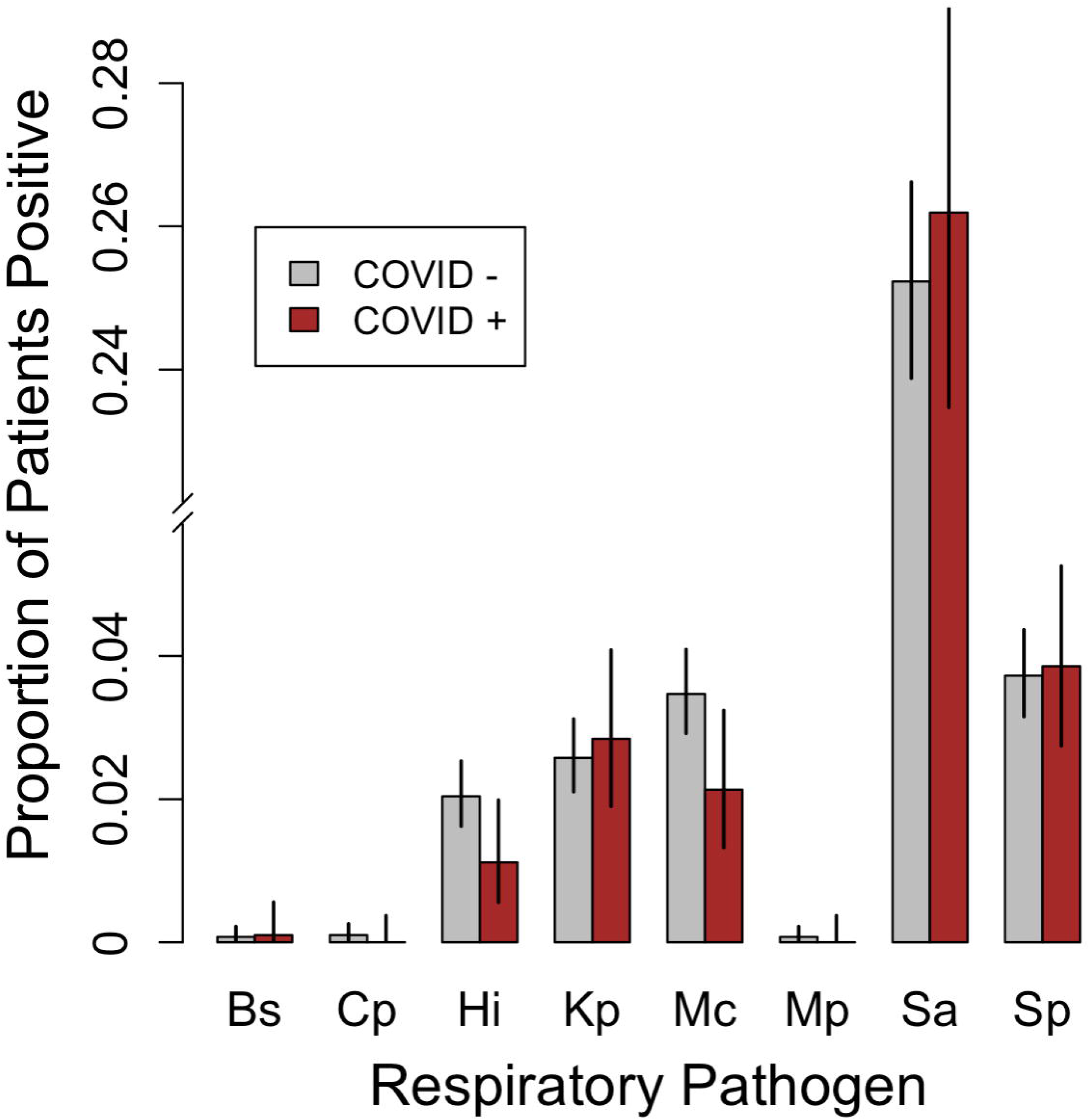
Grouped bar plot comparing the proportions of SARS-CoV-2 positive patients (n = 985) and SARS-CoV-2 negative patients (n = 3920) that tested positive for the various respiratory bacterial pathogens detected during the study period. Error bars represent 95% confidence intervals for the estimated proportions. Bs = *Bordetella* sp., Cp = C. *penumoniae*, Hi = *H. influenzae*, Kp = *K. pneumoniae*, Mc = *M. catarrhalis*, Mp = *M. pneumoniae*, Sa = S. aureus, Sp = S. pneumoniae. Note that *H. influenzae B, L. pneumophila*, and *Salmonella sp*. were not detected in any patients and were therefore omitted.

Of the 4905 patients included in this study 1566 (31.9%) tested positive for at least one bacterial pathogen and 471 (9.6%) patients tested positive for a pathogen other than *S. aureus*. Additionally, 213 (4.3%) patients tested positive for multiple bacterial pathogens and 85 (1.7%) patients tested positive for multiple pathogens not counting S. *aureus* (Table 2). Contingency tables, odds ratios, 95% confidence intervals, and *P*-values from Fisher’s Exact Tests assessing the association between SARS-CoV-2 and bacterial infection of any kind, bacterial infections other than S. *aureus*, polymicrobial infection including *S. aureus*, and polymicrobial infection not including *S. aureus* are shown in Table 2. Upon correcting for multiple testing, no significant associations were detected. However, the percentage of patients in which multiple non-*S*.*aureus* bacteria were detected was more than twice as high in SARS-CoV-2 negative patients (1.9%) when compared to SARS-CoV-2 positive (0.91%) patients (Table 2).

**Table 2.** Association between SARS-CoV-2 status and number of patients that tested positive for the various bacterial respiratory pathogens detected during the study period.

|  | COVID Status |  | OR (95% CI) | p-value | Adjusted p-value |
| --- | --- | --- | --- | --- | --- |
| **One or more Bacteria of any kind** | Positive | Negative |  |  |  |
| Yes | 309 | 1257 |  |  |  |
| No | 676 | 2663 |  |  |  |
| Percentage | 31 | 32 | 0.9684 (0.8302-1.1281) | 0.7023 | 0.9306 |
| **One or more bacteria excluding *S. aureus*** |  |  |  |  |  |
| Yes | 87 | 384 |  |  |  |
| No | 898 | 3536 |  |  |  |
| Percentage | 8.8 | 9.8 | 0.8921 (0.6907-1.1426) | 0.3970 | 0.7940 |
| **Multiple bacteria of any kind** |  |  |  |  |  |
| Yes | 42 | 171 |  |  |  |
| No | 943 | 3749 |  |  |  |
| Percentage | 4.3 | 4.4 | 0.9765 (0.6743-1.3871) | 0.9306 | 0.9306 |
| **Multiple bacteria excluding *S. aureus*** |  |  |  |  |  |
| Yes | 9 | 76 |  |  |  |
| No | 976 | 3844 |  |  |  |
| Percentage | 0.91 | 1.9 | 0.4665 (0.2047-0.9374) | 0.02806 | 0.11224 |

Raw counts, odds ratios (OR), 95% confidence intervals (95% CI), and p-values based on Fisher's Exact
Test of the null hypothesis that the true odds ratio is equal to one. Odds ratios describe how many times
more likely bacterial infection is given the presence of SARS-CoV-2.

## Discussion

Among patients infected with SARS-CoV-2 globally, meta-analyses report a wide range (1-100%) of antimicrobial prescribing rates. In the United States these rates generally range from 50% to 95% and are typically near 70%^16-19^. While some of these patients do suffer from bacterial co-infection or develop secondary infections after admission, a disparity may exist between the rate of prescribing antibiotics and the number of patients concurrently infected with SARS-CoV-2 and bacterial pathogens. As antibiotics are prescribed, adverse drug reactions and antimicrobial resistance become concerns. With studies identifying drug resistant organisms in SARS-CoV-2 patients treated with antibiotics, it has become imperative to determine the necessity of utilizing these agents^20,21^. With studies identifying drug resistant organisms in SARS-CoV-2 patients treated with antibiotics, it has become imperative to weigh the risks and benefits of utilizing these agents. To address the necessity of prescribing antibiotics during the current pandemic, we explored the propensity of SARS-CoV-2 infected patients to also experience bacterial co-infection in the state of Texas. Further, we investigated whether patient demographics were associated with the rate of SARS-CoV-2 or bacterial infection, as well as the types of bacteria and number of polymicrobial infections present.

The rate of co-infection varies widely in the literature ranging from approximately 1 to 50 percent^17,20,22-28^. Many factors may contribute to this variation including differences in geography, setting (inpatient vs outpatient), time of collection (upon admission vs during hospital stay), site of collection/sample type (sputum vs bronchiolar lavage vs nasopharyngeal swab etc.), and targeted organisms (bacteria vs viruses or both). This study detected bacteria in 31.9 percent of patients including 32 percent among SARS-CoV-2 positive patients and 31 percent among SARS-CoV-2 negative patients. While this bacterial co-infection rate may be higher than those estimated from many studies, it is typical within the literature for studies utilizing nasopharyngeal swabs to observe higher rates of bacteria compared to those drawing from other sources such as sputum. For example, a study conducted by Calcogno et al., in which 75.9% of samples were collected via nasopharyngeal swabs, reported a similar rate (32.7 percent of patients overall in comparison to 31.9 percent in our study)^25^. Additionally, Calcogno et al.^25^ identified *S. aureus* in 15.4 percent of patients while *S. aureus* was identified in 25.4% percent of patients in our study. When combined with the notion that *S. aureus* colonizes the nares of a large portion of the general population, it’s likely that the rate of infection, especially that by *S. aureus* was inflated due to commensal bacteria. In addition to S. *aureus* our study identified *H. influenza* (1.8%), *K. pneumoniae* (2.6%), *S. pneumoniae* (3.8%) and *M. catarrhalis* (3.2%) as the most common pathogens. Similarly, in a meta-analysis by Musuuza et al., the most common respiratory pathogens detected in patients co-infected with SARS-CoV-2 were K. *pneumoniae* (9.9%), *S. pneumoniae* (8.2%), *S. aureus* (7.7%). *H. influenzae* (6.6%), and *M. pneumoniae* (4.3%), and M. catarrhalis (1.7%) ^29^. Unlike in our study however, Musuuza et al.^29^ found that *M. pneumoniae* comprised 4.3% of the bacterial pathogens identified, whereas our study only detected three instances of *M. pneumoniae*, all of which were SARS-CoV-2 negative patients.

While many factors potentially contribute to the similarities and differences between our results and those of others, it is important to note that the types of bacterial infections observed may change over time and/or differ considerably between outpatient and hospital settings. For example, *Acinetobacter, Pseudomonas*, and *Enterococcus* were more commonly identified after prolonged hospital stays and multi-drug resistant organisms (MDROs) have been detected in hospitalized patients^20,29^. An additional limitation of comparing our results to the results of others is that the panels of bacterial pathogens tested for often differs among studies. Thus, while it is expected for pathogens associated with prolonged hospital stays to be less frequent in our outpatient population, use of the Fast Tracking Diagnostics (FTD) panel limited our ability to fully assess their impact because it does not target many of these organisms. Nevertheless, while the FTD panel may have limited the number of bacterial species detected and therefore the overall infection rate, this limitation was partially addressed by noting the presence of individual species. Additionally, separate analyses were conducted with and without S. *aureus*. In doing so, infection rates of specific pathogens could be compared across groups or between studies. Despite these limitations, this study is unique in that few studies have specifically reviewed outpatient data and while several studies described the rate of co-infection in COVID-19 patients, few studied the difference in rate among SARS-CoV-2 positive and negative patients. However, due to a lack of clinical follow up, culture, or genomic information, we were not able to describe the effect of antimicrobial treatment on these patients. Overall, this study, and comparisons of our data with data from other studies, show no evidence of an increased rate of bacterial infection among SARS-CoV-2 positive patients.

In addition to comparing the rate and types of bacterial infections in SARS-CoV-2 positive and negative patients we sought to identify populations that may be at increased risk of either SARS-CoV-2 or bacterial infections. The data from this study confirm an increased rate of SARS-CoV-2 present in Hispanics. This study was limited by its retrospective nature and self-reported demographics, of which many patients declined to answer. Despite this limitation the proportion of Hispanics testing positive for SARS-CoV-2 is in line with data reported by the CDC^30^. Notably 2639 patients did not disclose their ethnicity and only 283 patients identified as non-Hispanic compared to 1983 patient who identified as Hispanic. Consequently, the ethnicity of 53.8% of the patients is unknown and therefore volunteer bias should be considered when interpreting these results. Despite this limitation, this study did uncover that Hispanics were significantly more likely to be infected with SARS-CoV-2 when compared to non-Hispanics or when compared to those that did not self-report their ethnicity. Others have reported similar observations, suggesting minorities have higher incidence and mortality rates due to COVID-19 ^31-33^. This study also found that *S. aureus* was more likely to be observed in men and adults below the age of 65, while other patient demographics did not appear to influence the presence of bacteria.

RT-PCR is currently the gold standard for detecting SARS-CoV-2, and in this study, this technology was used concurrently with RT-PCR-based bacterial pathogen testing to ensure that no systemic discrepancies existed in the sample collection and testing procedures. Using this approach, we found a higher rate of detection of SARS-CoV-2 in Hispanics; however, no significant differences were detected in the rate of bacterial infection between SARS-CoV-2 positive and SARS-CoV-2 negative patients. Therefore, these data alone do not support an increase in the prescription of antimicrobials due to the presence of SARS-CoV-2. We also detected *S. aureus* at higher rates in men and people under 65. However, *S. aureus* (the most commonly detected bacterium in our study) is observed in the nares of approximately 20%-30% of the population^34,35^, suggesting that the presence of *S. aureus* in and of itself is not necessarily indicative on infection—an issue we addressed by conducting separate analyses that included and excluded *S. aureus*. Because this study was derived from outpatient settings at multiple sites within the state of Texas, these data may not generalize to populations from other parts of the US. However, by surveying outpatients, this study may be more applicable to the portion of the general public that does not require hospitalization. Moreover, this study may provide utility in selecting appropriate empiric therapies for patients who develop serious infections that require hospitalization.

## Methods

### Study design, sample collection & patient population

This study was a retrospective, multicenter analysis of outpatients concurrently tested for the presence of SARS-CoV-2 as well as respiratory pathogens from April 2020 to April 2021. The patient population was derived from within the state of Texas and patients were indicated for testing due to experiencing symptoms such as cough, fever, etc.

Sample kits composed of requisition forms, nasopharyngeal swabs (Huachenyang iClean, Shenzhen, China) and conical tubes (Stellar Scientific, Baltimore Maryland) containing storage buffer were sent to outpatient facilities. Storage buffer was prepared according to CDC guidleines^36^. Patient demographics including sex, date of birth, ethnicity and race were included on the requisition forms and self-reported by patients. Broad consent was provided by the patients at the time of testing. Prior to analysis patient information was de-identified using the Safe Harbor Method^37^. Specifically, date of birth, provider, facility, insurance provider, and detailed geographic information such as zip codes etc. were removed from the data set. This study was exempted from institutional review by the Institutional Review Board at Texas A&M University-San Antonio (TAMUSA IRB #2022-016).

### RNA isolation and Pathogen Identification

Bacteria were characterized by species, using RT-PCR. After nasal swab samples were obtained, RNA was extracted using the Applied Biosystems MagMax Viral/Pathogen II kit (ThermoFisher, Waltham, MA) per the manufacturer’s instructions. After isolation, RNA was amplified and detected via RT-PCR using the Applied Biosystems QuantStudio 7 flex and 12K flex systems (ThermoFisher, Waltham, MA). Primers, probes and enzymes used to detect SARS-CoV-2 and bacteria were purchased from Integrated DNA Technologies (Coralville, Iowa) and Fast Track Diagnostics (Sliema, Malta) respectively. Positive SARS-CoV-2 tests were defined as a cycle threshold (Ct) value of 40 or less for RNAse P, N1 and N2 collectively. The Fast Track Diagnostics (FTD) respiratory kit contained probes for *Bordetella spp, Chlamydia pneumoniae, Haemophilus influenzae, Haemophilus influenzae type B, Klebsiella pneumoniae, Legionella pneumophila, Moraxella catarrhalis, Mycoplasma pneumoniae, Salmonella spp, Staphylococcus aureus, and Streptococcus pneumoniae*. Bacterial infection was defined as a positive RT-PCR test in which the Ct value was less than 35 for any of the respective probes.

### Statistical analysis

This study is based on a subset of a larger data set generated from over 20 states and containing more than 28,000 SARS-CoV-2 tests. These tests were conducted between the months of April 2020 and April 2021. All patients included in this study were administered SARS-CoV-2 and respiratory pathogen tests concurrently. To ensure that analyses were based on a common suite of tests, only patients tested for SARS-CoV-2 and all 11 respiratory pathogens described above were included. Additionally, to ensure that all observations were independent, patients that were continually monitored or otherwise received multiple tests were excluded. Given the differences in mitigation policy between states, and the fact that Texas was the most heavily represented state in this data set, we focused on patients residing in Texas for this study. After imposing these filters, the analyses we conducted were based on 4905 patients in total. These data are composed of patients from throughout the state of Texas; however, 1522 (31%) samples (i.e., patients) were collected in the McAllen-Edinburgh-Mission metropolitan statistical area (MSA) and 2357 (48%) samples were collected from the San Antonio-New Braunfels MSA. Thus, 79% of the samples that our analyses are based on come from these two MSAs. In addition, these data include 1026 patients who did not disclose their city of residence, were from MSAs (e.g., Dallas-Fort Worth) other than McAllen-Edinburgh-Mission and San Antonio-New Braunfels, or rural areas throughout the state.

We used the ‘binom.test’ function in R to calculate point estimates and 95% confidence intervals for the proportion of patients positive for SARS-CoV2, S. *aureus*, and bacterial pathogens other than S. *aureus*. This approach was applied across several demographic categories and was complemented by using R to test for equal proportions (‘prop.test’ function) among: (1) races, (2) ethnicities, (3) women vs. men, and (4) patients ≥ 65 vs. patients < 65. We then adjusted the 12 resulting P-values (three pathogen categories x four demographic factors) for multiple testing using the False Discovery Rate (FDR = 0.05) correction described by Benjamini and Hochberg^38^.

In addition to examining positivity rates across demographic factors, we also used ‘binom.test’ to compute point estimates and 95% confidence intervals of the proportion of SARS-CoV-2 positive and SARS-CoV-2 negative patients who tested positive for each of the eight bacterial pathogens detected in our survey. We then used ‘prop.test’ to make comparisons between the bacterial positivity rates of SARS-CoV-2-positive and SARS-CoV-2-negative patients for bacterial pathogens that had sufficient data to ensure that all expected cell counts were five or larger. The resulting five *P*-values were adjusted for multiple testing by controlling the FDR at the 0.05 level^38^.

Finally, we used the odds ratio (*OR*) to estimate the magnitude and direction of association between: (1) SARS-CoV-2 and detection of bacterial pathogens of any kind, (2) SARS-CoV-2 and detection of bacteria other than *S. aureus*, (3) SARS-CoV-2 and detection of more than one bacterium of any kind, and (4) SARS-CoV-2 and detection of more than one kind of bacteria, not including *S. aureus*. The null hypothesis that *OR* = 1.00 was assessed via Fisher’s Exact Test and 95% confidence interval of *OR* were computed using the ‘fisher.test’ function in R. The four resulting p-values were corrected for multiple testing by controlling the FDR at the 0.05 level^38^.

## Data Availability

All data produced in the present study are available upon reasonable request to the authors.

## Author Contributions

Conceptualization, JFS, CAM, PKM, NV, and RBP. Methodology, JFS, RBP, PKM, KL, and VN. Analysis, JFS, RBP, CAM, VN, KL, PKM. Writing, revising and editing, JFS, CAM, RBP, NV, and PKM.

## Data Availability Statement

The data that support the findings of this study are available from the corresponding author, Dr. Pramod K. Mishra, upon reasonable request.

