## Supplemental Figure 1 for "Prevalence and Characterization of Bacteria Present in the Nasopharynx of Outpatients with and without SARS-CoV-2"

| **Demographic Factor** | **Pathogen** | | **Chi-Squared_Value** | **Degrees of Freedom** | **Unadjusted P-value** | **Adjusted P-value** | **Remarks** |
| --- | --- | --- | --- | --- | --- | --- | --- |
| Race (Asian, Black, White) | | SARS-CoV2 | 2.8533 | 2 | 0.2401 | 0.3373 | Not enough American Indians or Pacific Islanders to include in analysis |
| Race (Asian, Black, White) | | *S. aureus* | 0.13907 | 2 | 0.9328 | 0.9328 | Not enough American Indians or Pacific Islanders to include in analysis |
| Race (Black, White) | | Bacteria other than *S. aureus* | 0.019204 | 1 | 0.8898 | 0.9328 | Not enough Asian, American Indians or Pacific Islanders to include in analysis |
| Ethnicity (Hispanic, Not Hispanic | | SARS-CoV2 | 15.346 | 1 | 8.95E-05 | 0.0005 | NA |
| Ethnicity (Hispanic, Not Hispanic | | *S. aureus* | 4.1993 | 1 | 0.04044 | 0.1213 | NA |
| Ethnicity (Hispanic, Not Hispanic | | Bacteria other than *S. aureus* | 0.85158 | 1 | 0.3561 | 0.4273 | NA |
| Sex | | SARS-CoV2 | 1.9347 | 1 | 0.1643 | 0.3286 | NA |
| Sex | | *S. aureus* | 16.828 | 1 | 4.09E-05 | 0.0005 | NA |
| Sex | | Bacteria other than *S. aureus* | 1.6969 | 1 | 0.1927 | 0.3303 | NA |
| Age (>=65, <65) | | SARS-CoV2 | 1.3216 | 1 | 0.2503 | 0.3337 | NA |
| Age (>=65, <65) | | *S. aureus* | 9.51 | 1 | 0.002044 | 0.0082 | NA |
| Age (>=65, <65) | | Bacteria other than *S. aureus* | 3.3568 | 1 | 0.06693 | 0.1606 | NA |

| **Bacterial Pathogen** | **Chi-Squared_Value** | **Degrees of Freedom** | **Unadjusted P-value** | **Adjusted P-value** | **Remarks** |
| --- | --- | --- | --- | --- | --- |
| *Bordetella spp.* | NA | NA | NA | NA | Only deteted in 4 patients, not enough data to analyze |
| *C. pneumoniae* | NA | NA | NA | NA | Only detected in 4 patients, not enough data to analyze |
| *H. influenzae* | 3.2016 | 1 | 0.07357 | 0.1839 | NA |
| *K. pneumoniae* | 0.12617 | 1 | 0.7224 | 0.903 | NA |
| *M. catarrhalis* | 4.123 | 1 | 0.0423 | 0.1839 | NA |
| *M. pneumoniae* | NA | NA | NA | NA | Only detected in 3 patients, not enough data to analyze |
| *S. aureus* | 0.33615 | 1 | 0.5621 | 0.903 | NA |
| *S. pneumoniae* | 0.010641 | 1 | 0.9178 | 0.9178 | NA |
